# Severe hospital events following symptomatic infection with Sars-CoV-2 Omicron and Delta variants in France, December 2021 – January 2022: a retrospective, population-based, matched cohort study

**DOI:** 10.1101/2022.02.02.22269952

**Authors:** Vincent Auvigne, Sophie Vaux, Yann Le Strat, Justine Schaeffer, Lucie Fournier, Cynthia Tamandjou, Charline Montagnat, Bruno Coignard, Daniel Levy-Bruhl, Isabelle Parent du Châtelet

**Affiliations:** Santé publique France - 12 rue du Val d’Osne - 94415 Saint-Maurice - France; Sully - 3, av. Doyen Louis Weil - 38000 Grenoble - France

## Abstract

**Background:** A rapid increase in incidence of the SARS-CoV-2 Omicron variant occurred in France in December 2021, while the Delta variant was prevailing since July 2021. We aimed to determine whether the risk of a severe hospital event following symptomatic SARS-CoV-2 infection differs for Omicron versus Delta.

**Methods:** We conducted a retrospective cohort study to compare severe hospital events (admission to intensive care unit or death) between Omicron and Delta symptomatic cases matched according to week of virological diagnosis and age. The analysis was adjusted for age, sex, vaccination status, presence of comorbidities and region of residence, using Cox proportional hazards model.

**Findings:** Between 06/12/2021-28/01/2022, 184 364 cases were included, of which 931 had a severe hospital event (822 Delta, 109 Omicron). The risk of severe event was lower among Omicron versus Delta cases; the difference in severity between the two variants decreased with age (aHR=0·11 95%CI: 0·07-0·17 among 40-64 years, aHR=0·51 95%CI: 0·26-1·01 among 80+ years). The risk of severe event increased with the presence of comorbidities (for very-high-risk comorbidity, aHR=4·18 95%CI: 2·88-6·06 among 40-64 years) and in males (aHR=2·29 95%CI: 1·83-2·86 among 40-64 years) and was higher in unvaccinated compared to primo-vaccinated (aHR=6·90 95%CI: 5·26-9·05 among 40-64 years). A booster dose reduced the risk of severe hospital event in 80+ years infected with Omicron (aHR=0·27; 95%CI: 0·11-0·65).

**Interpretation:** This study confirms the lower severity of Omicron compared to Delta. However, the difference in disease severity is less marked in the elderly.

## Introduction

The SARS-CoV-2 Omicron variant (B1.1.529, BA.*) was first reported on November 23 2021, in Gauteng Province, South Africa.^1^ The World Health Organization ^2^ designated Omicron as a variant of concern (VOC) due to the presence of more than 30 mutations in the spike gene of the virus, its greater transmissibility, an increased risk of reinfection and its potential immune escape. As in many other European countries, sequencing data have shown a rapid increase of the Omicron variant in France: it represented 10.7% of interpretable sequences on December 13, 2021, almost 50% on December 20, 2021 and 89% during the first week of 2022. As of January 3, 2022, the Omicron variant has been detected in all regions of France.^3,4^ The overall risk of severe disease associated to this variant appears to be lower than Delta-related cases. In South Africa, descriptive analyses were performed to compare the Omicron-dominant wave to previous SARS-CoV-2 waves. The authors observed fewer hospitalisations, less severe hospitalized cases, and a shorter length of hospital stay (3 days versus 7-8 days) during the Omicron wave.^5-9^ However, the observed lower number of severe events among Omicron cases could reflect changes over time in the characteristics of the infected population. The primary objective of this study was to determine whether the risk of occurrence of a severe hospital event in adults following symptomatic SARS-CoV-2 infection differs for Omicron versus Delta. Secondary objectives were to assess whether this risk differs with age, sex, vaccination status and comorbidities.

## Methods

### Study design

This is a retrospective cohort study carried out by combining three French COVID-19 national surveillance databases. The study covers the period from 06/12/2021 to 28/01/2022, during which the Omicron and Delta variants co-circulated in France. The outcome of the study was the occurrence of a severe hospital event associated with COVID-19 i.e. admission to intensive care unit (ICU) or death in hospital, among symptomatic persons testing positive for SARS-CoV-2, classified as Delta or Omicron using mutation screening. Delta and Omicron cases were matched according to age and calendar week of virological diagnosis. Cox proportional hazards model was used to estimate the adjusted hazard ratio (aHR); the models were adjusted for age, sex, vaccination status, presence of comorbidity and region of residence.

### Data sources

We used pseudonymised health data from three information systems. These three databases respectively include results of all SARS-CoV-2 tests (RT-PCR and antigen tests, excluding self-tests) performed in the country (SI-DEP), summary data on hospital stays for COVID-19, outcomes and types of wards (SI-VIC) and COVID-19 vaccine status and comorbidities of the French population (VAC-SI). Data were extracted on 01/02/2022. As four days are required for 90% of hospital events to be reported in the SI-VIC database, events after 28/01/2022 were excluded to control for reporting bias. A pseudonym, resulting from a cryptographic hash based on surname, first name, date of birth and sex was used to merge the three data sources.

### Eligibility criteria

Persons ≥18 years old on the day of inclusion, sampled for SARS-CoV-2 diagnostic between 06/12/2021 and 23/01/2022 (inclusive) and declaring themselves as symptomatic at sampling, with a confirmed SARS-CoV-2 infection and valid RT-PCR mutation screening were included. Exposed persons are those with a strong suspicion of infection with Omicron, based on the mutation screening results (see “Variant” below). Non-exposed persons are those with a strong suspicion of infection with Delta. Persons with missing data, as well as those not found in the VAC-SI database or with an inconsistent vaccination status were excluded.

### Variables

#### Virological analysis

For persons with several positive RT-PCR tests and valid mutation screening results, the first one was selected. If another positive test (RT-PCR or antigen test) without mutation screening result had been performed in the 15 days preceding the date of the RT-PCR test with mutation screening, the date of inclusion in the cohort was the date of this first test.

#### Variant

Characterization of samples that tested positive for SARS-CoV-2 by RT-qPCR was performed using a mutation screening multiplex RT-qPCR targeting a set of predefined mutations. During the study period, the screening strategy included three individual mutations (E484K, E484Q and L452R) and a panel of mutations associated with Omicron (deletion 69-70 or substitutions K417N, N501Y, S371L-S373P or Q493R). The detection of L452R was used as a proxy for Delta. The absence of E484K and L452R or the detection of one of the mutations associated with Omicron was used as a proxy for Omicron. To validate this algorithm, a study was performed on 11,574 results with sampling date between 01/12/2021 and 10/01/2022 for which a mutation screening result and an interpretable sequencing result were available. The specificity of the classification by the mutation screening was estimated at 99.1% for Delta and 92.9% for Omicron. (See supplementary material).

#### Vaccination status

At the date of inclusion, participants were classified as: unvaccinated, vaccinated with a complete primary vaccination or vaccinated with primary vaccination and a booster. Following French recommendations, the number of vaccine doses corresponding to each vaccination status depended also on the history of infection and immunosuppression. Doses given less than 14 days before the date of inclusion in the cohort, for the first dose, and less than 7 days for the subsequent ones were not retained. Persons with an incomplete primary vaccination schedule were considered unvaccinated.

#### Comorbidities

Comorbidities associated with complications are reported in VAC-SI as very-high-risk or medium-risk comorbidities. Very-high-risk comorbidities included cancers, haematological malignancies undergoing chemotherapy, severe chronic kidney disease, chronic dialysis, solid organ transplants, haematopoietic stem cell allografts, chronic multi-disease conditions with two or more organ failures, certain rare diseases and those at particular risk of infection, and Down’s syndrome.^10^ Medium-risk comorbidities included obesity, diabetes, chronic renal failure, chronic obstructive pulmonary disease, respiratory failure, hypertension, heart failure.

#### Hospital events

A person was considered to have had a “severe hospital event”, if he or she was, during the study period, admitted to an ICU or died in hospital without being admitted to an ICU. People tested at the hospital on the day of hospitalisation were included in the study. Only hospital events stated “for Covid” in the SI-VIC database (as opposed to “with Covid”, i.e. a person tested positive for SARS-CoV-2 but was hospitalised for a reason other than COVID-19) were included in the study.

### Matching

Each Omicron case was individually matched with a Delta case of the same age range (18-39, 40-64, 65-79, 80+) and with the same week of virological diagnosis. Unmatched individuals were excluded.

### Statistical methods

Kaplan-Meier curves were used to describe the temporal distribution of the outcome and the evolution of the cohort over the follow-up period. Univariate and multivariate analyses were performed using Cox proportional hazards model, taking into account individual matching. If the outcome occurred on the entry date, the follow-up time was considered 0·5 days. The multivariate models included the variables selected from the univariate analyses (p <0.2). Interactions were tested between variants and age, sex, vaccination status and comorbidities, and between age and sex, vaccination status and comorbidities. The effect of the time between the last vaccine dose and the virological diagnosis was tested. The multivariate models were constructed through a two-way stepwise selection process based on the Akaike information criterion (AIC). The analyses were performed using R. 4.0.4

## Results

### Participants

The number of individuals at different stages of the inclusion process are as follows:

- People over 18 with symptoms, sample taken during the inclusion period and RT-PCR positive with a valid result for mutation screening: 932 137
- Exclusion of 20 575 persons with missing sex or postal code in SIDEP: remaining 911 562
- Exclusion of 136 866 persons not found in VAC-SI: remaining 774 696
- Matching: 92 182 pairs constituted (184 364 participants) and exclusion of unmatched persons

### Descriptive data

Weekly numbers in the two groups of the cohort reflected the dynamic of the spread of the Omicron variant in France during December 2021 (Table 1). The majority (83%) of inclusions took place between weeks 2021-51 and 2022-01, as few Omicron cases were observed before and few Delta cases after. The proportion of unvaccinated individuals was more than twice as high in the Delta group as in the Omicron group (35·2% vs. 16·9%). Overall, 84% of first vaccine doses were with Pfizer/BioNTech vaccine, 9·8% with Moderna, 4·8% with AstraZeneca and 1·2% with Janssen, with no difference between the two groups. The proportion of people who have had a booster shot, and the median time between the booster shot and the virological diagnosis increased with age (Table 2), reflecting the phased roll-out of vaccination in France since the beginning of 2021.

**Table 1.**
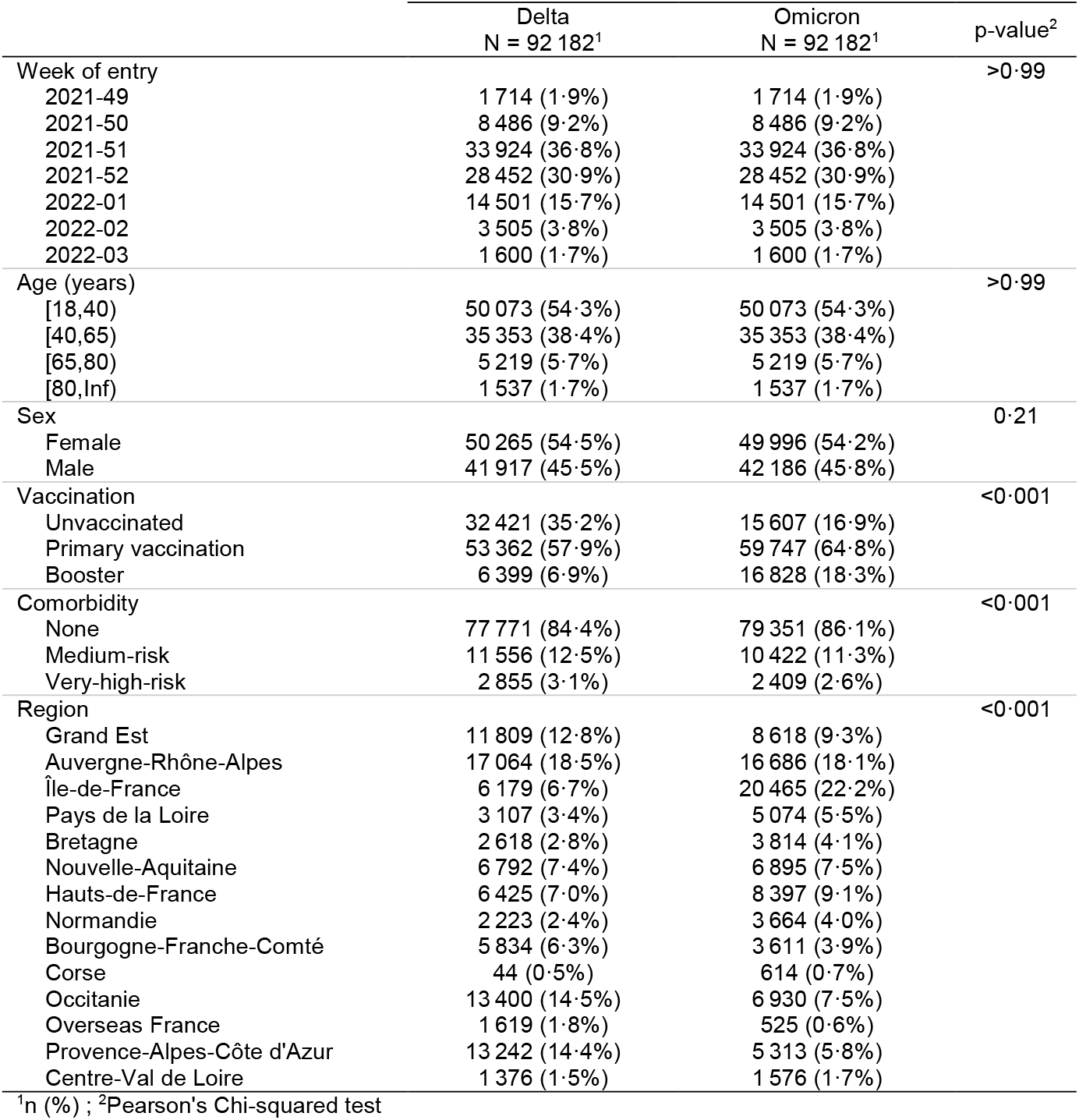
Baseline characteristics of participants at time of entry into study

**Table 2.** Vaccination status by age group, December 2021, France, (n= 184 364)

|  | [18,40)<br>N = 100 146<sup>1</sup> | [40,65)<br>N = 70 706<sup>1</sup> | [65,80)<br>N = 10 438<sup>1</sup> | [80,Inf)<br>N = 3 074<sup>1</sup> | p-value<sup>2</sup> |
| --- | --- | --- | --- | --- | --- |
| Unvaccinated | 27 281 (27 %) | 17 414 (25%) | 2 465 (24%) | 868 (28%) | <0.001 |
| Primary vaccination | 66 060 (66%) | 42 433 (60%) | 3 773 (36%) | 843 (27%) |  |
| Booster | 6 805 (6.8%) | 10 859 (15%) | 4 200 (40%) | 1 363 (44%) |  |
| Days since booster<sup>3</sup> | 17 (10, 27) | 20 (11, 31) | 36 (21, 58) | 69 (37, 96) |  |

<sup>1</sup>n (%); <sup>2</sup>Pearson's Chi-squared test; <sup>3</sup>Median, Inter Quartile Range

### Follow-up and outcome data

The maximum follow-up duration was 53 days; 94% of participants were followed up for at least 20 days, or had a severe hospital event before that time point (Figure 1). A total of 931 severe hospital events were observed, including 699 ICU admissions and 232 deaths without ICU admission. The number of severe hospital events was significantly higher (p<0·001) in the Delta group (822) than in the Omicron group (109). In the Delta group, 82% of severe events were observed within 10 days of positive sampling, and 96% within 20 days. In the Omicron group, 68% of events occurred within 10 days and 88% within 20 days (Figure 1).

**Figure 1.**
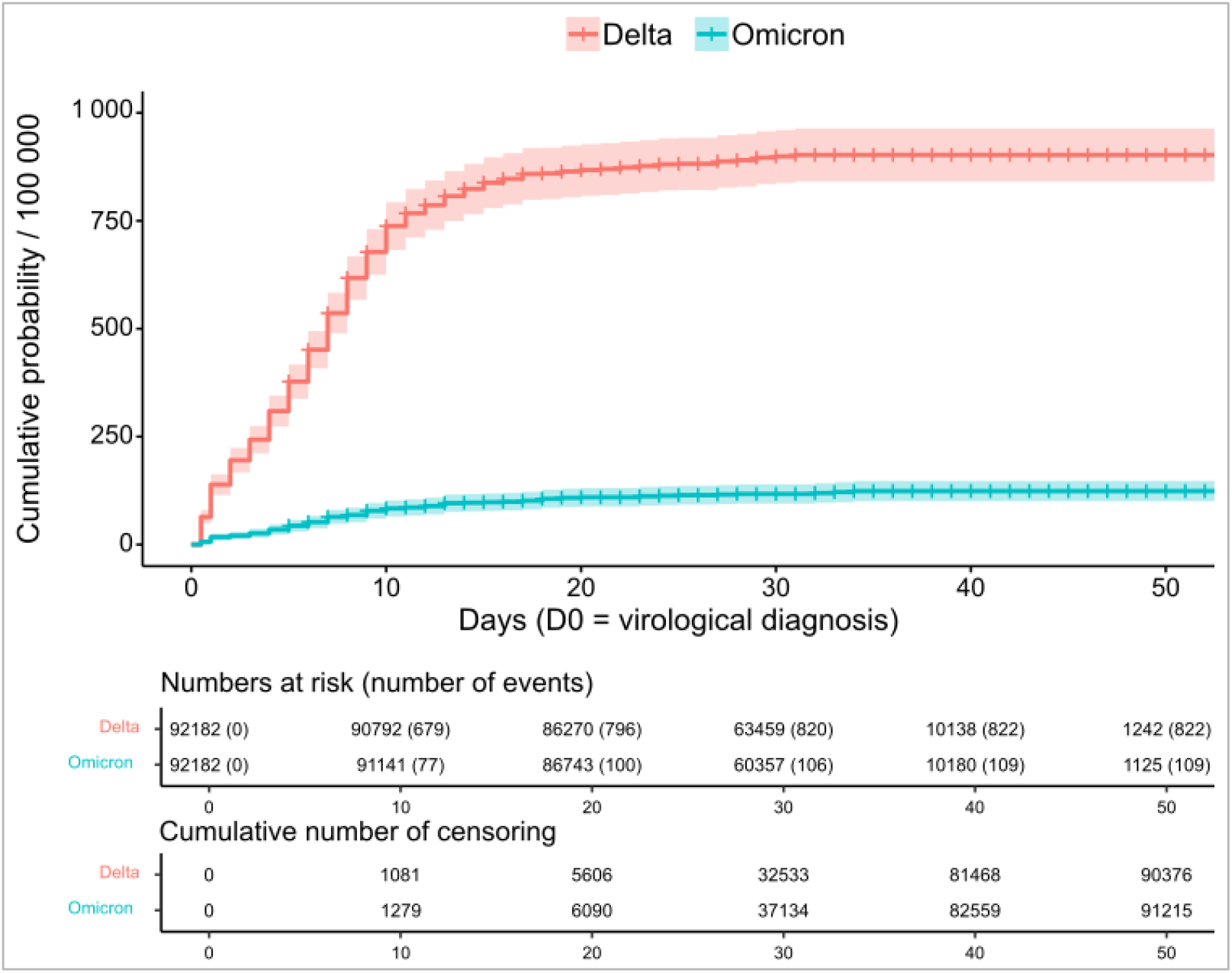
Cumulative probability of a severe hospital event among cases infected by the Omicron or Delta variant of the SARS-CoV-2 by time since virological diagnosis, December 2021-January 2022, France, (n= 184 364)

### Univariate analysis

Table 3 gives, for each covariate value, the crude hazard ratio ^11^ of a severe hospital event and the absolute number of severe events observed per 100 000 people within 20 days of positive sampling. The cHR was statistically different between the two groups of the cohort for all covariate modalities (p<0·05), except for the region (statistically significant for 6 of the 13 regions).

**Table 3.**
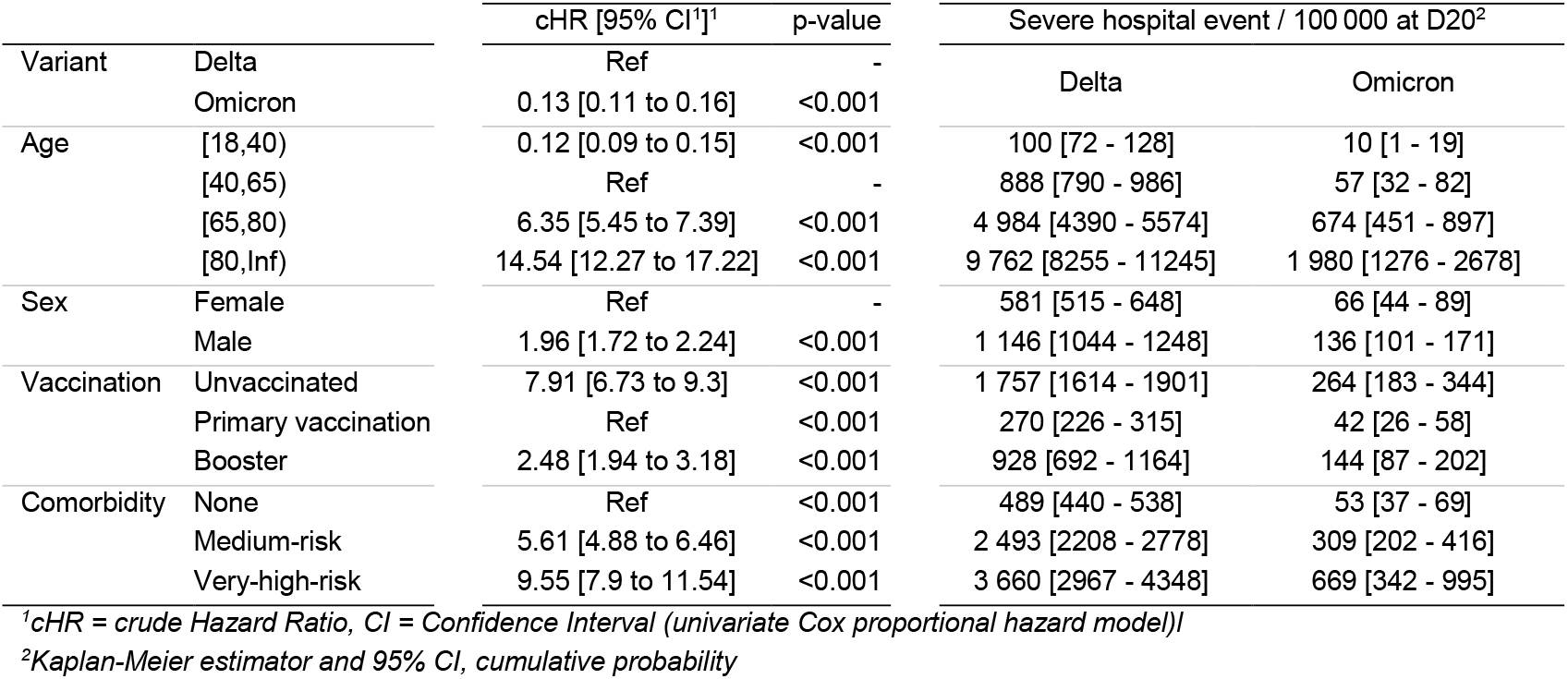
Risk of a severe hospital event among cases infected by the Omicron or Delta variant of the SARS-CoV-2 (univariate analysis), December 2021-January 2022, France (n= 184 364)

### Multivariate analysis

All the variables were included in the multivariate analyses. Models were adjusted for region of residence (not shown). The overall model showed a risk reduction in the Omicron group, a drastic increase of the risk with age, but also interactions between variant and age, age and vaccination, age and comorbidity (Table 4). Consequently, the models were stratified by age group (Table 5). The 80+ model showed an interaction between variant and vaccination; therefore, this age group was stratified by variant (Table 6). The risk of having a severe hospital event is lower in the Omicron group, in each age group; the difference in severity between the two variants decreased with age. People aged 40-64 years with Omicron-related symptomatic infection had a 9·1 times lower risk (5·9-14·3) of having a severe hospital event than those with Delta-related symptomatic infection [aHR=0.11; 95% CI: 0·07 to 0·17]. In people aged 80 years and over, the Omicron cases had only a 2-fold lower risk (1 to 3·8) than Delta cases [aHR=0.51; 95% CI: 0·26-1·01]. The models also highlighted the magnitude of the risk associated with other covariates. The risk increased with the presence of comorbidity; the risk associated with very-high-risk comorbidities being higher than the risk associated with medium-risk comorbidities and the risk associated with comorbidities decreasing with age. The risk was almost twice as high for men as for women, with no major difference between age groups.

**Table 4.** Risk of a severe hospital event among cases infected by the Omicron or Delta variant of the SARS-CoV-2 (multivariate analysis), overall model with interactions, December 2021-January 2022, France (n= 184 364)

|  | aHR (95% CI)<sup>1</sup> | p-value |
| --- | --- | --- |
| **Variant** |  |  |
| DELTA | Ref |  |
| OMICRON | 0.10 (0.07 to 0.16) | <0.001 |
| **Age** |  |  |
| [18,40) | 0.09 (0.04 to 0.19) | <0.001 |
| [40,65) | Ref |  |
| [65,80) | 9.76 (6.84 to 13.9) | <0.001 |
| [80,Inf) | 30.9 (20.0 to 47.7) | <0.001 |
| **Sex** |  |  |
| Female | Ref |  |
| Male | 2.12 (1.86 to 2.42) | <0.001 |
| **Vaccination** |  |  |
| Unvaccinated | 7.14 (5.45 to 9.35) | <0.001 |
| Primary vaccination | Ref |  |
| Booster | 1.04 (0.58 to 1.88) | 0.90 |
| **Comorbidity** |  |  |
| None | Ref |  |
| Medium-risk | 3.03 (2.40 to 3.82) | <0.001 |
| Very-high-risk | 4.07 (2.81 to 5.90) | <0.001 |
| **Variant \* Age** |  |  |
| OMICRON * [18,40) | 1.59 (0.61 to 4.10) | 0.34 |
| OMICRON * [65,80) | 1.94 (1.13 to 3.32) | 0.016 |
| OMICRON * [80,Inf) | 3.14 (1.78 to 5.54) | <0.001 |
| **Age \* Vaccination** |  |  |
| [18,40) * Unvaccinated | 1.46 (0.65 to 3.27) | 0.36 |
| [65,80) * Unvaccinated | 0.54 (0.37 to 0.79) | 0.001 |
| [80,Inf) * Unvaccinated | 0.40 (0.26 to 0.62) | <0.001 |
| [18,40) * Booster | 2.95 (0.57 to 15.4) | 0.20 |
| [65,80) * Booster | 0.81 (0.40 to 1.63) | 0.55 |
| [80,Inf) * Booster | 0.53 (0.24 to 1.13) | 0.10 |
| **Age \* Comorbidity** |  |  |
| [18,40) * Medium-risk | 0.88 (0.42 to 1.82) | 0.73 |
| [65,80) * Medium-risk | 0.57 (0.40 to 0.79) | <0.001 |
| [80,Inf) * Medium-risk | 0.43 (0.29 to 0.62) | <0.001 |
| [18,40) * Very-high-risk | 1.58 (0.53 to 4.75) | 0.41 |
| [65,80) * Very-high-risk | 0.74 (0.46 to 1.18) | 0.21 |
| [80,Inf) * Very-high-risk | 0.40 (0.24 to 0.69) | <0.001 |

<sup>1</sup>aHR = adjusted Hazard Ratio, CI = Confidence Interval

**Table 5.**
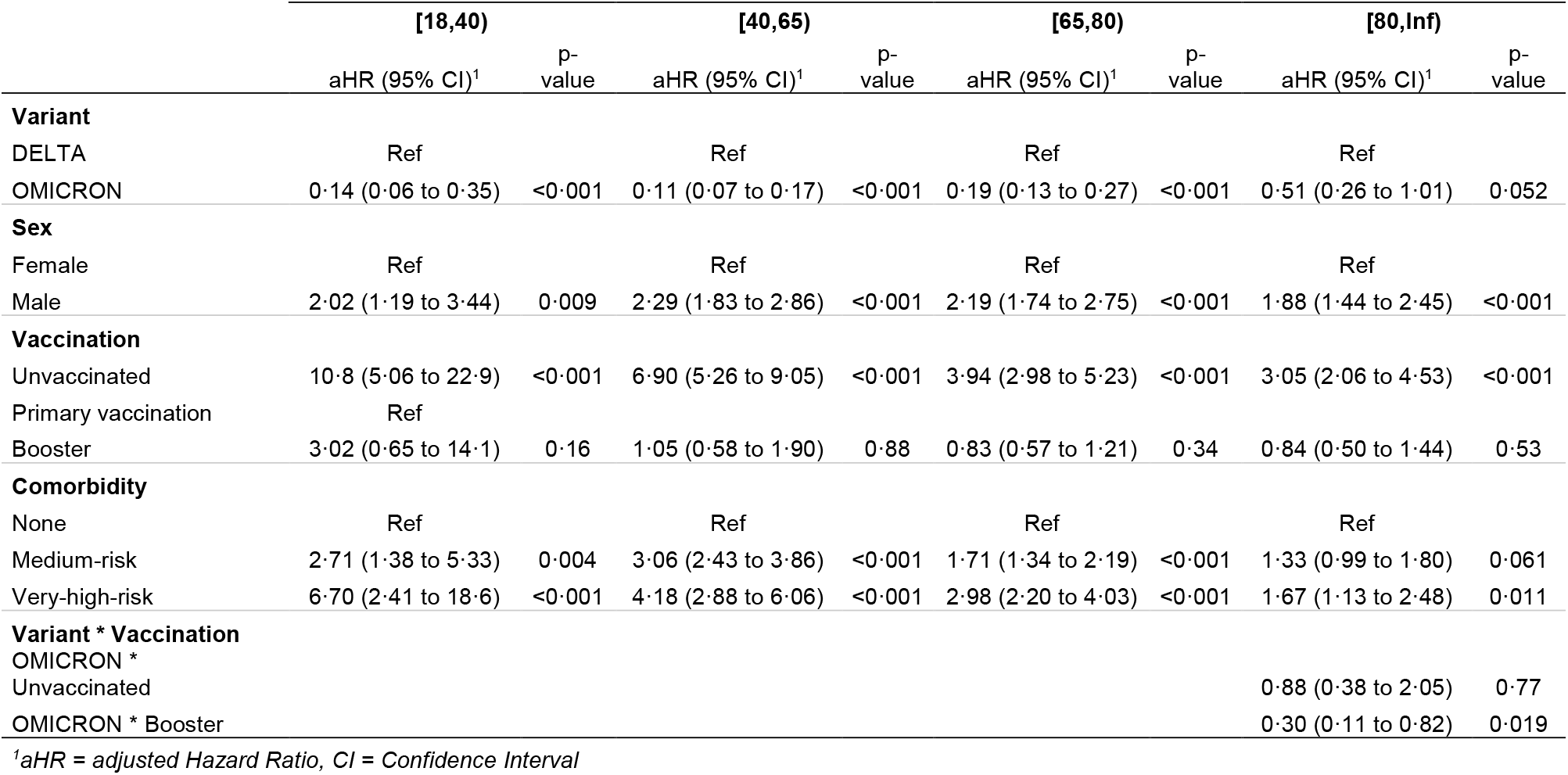
Risk of a severe hospital event among cases infected by the Omicron or Delta variant of the SARS-CoV-2, stratified per age (multivariate analysis), December 2021-January 2022, France (n= 184 364)

**Table 6.** Risk of a severe hospital event among cases infected by the Omicron or Delta variant of the SARS-CoV-2, 80 years and above, stratified per variant (multivariate analysis), December 2021-January 2022, France (n= 3 074)

|  | Delta |  | Omicron |  |
| --- | --- | --- | --- | --- |
|  | aHR (95% CI)<sup>1</sup> | p-value | aHR (95% CI)<sup>1</sup> | p-value |
| **Sex** |  |  |  |  |
| F | Ref |  | Ref |  |
| M | 1.96 (1.45 to 2.64) | <0.001 | 1.50 (0.79 to 2.84) | 0.22 |
| **Vaccination** |  |  |  |  |
| Unvaccinated | 3.07 (2.07 to 4.57) | <0.001 | 2.69 (1.28 to 5.64) | 0.009 |
| Primary vaccination | Ref |  | Ref |  |
| Booster | 0.82 (0.48 to 1.40) | 0.47 | 0.27 (0.11 to 0.65) | 0.003 |
| **Comorbidity** |  |  |  |  |
| None | Ref |  | Ref |  |
| Medium-risk | 1.27 (0.91 to 1.78) | 0.17 | 1.60 (0.82 to 3.12) | 0.17 |
| Very-high-risk | 1.95 (1.28 to 2.98) | 0.002 | 0.46 (0.10 to 2.06) | 0.31 |

<sup>1</sup>aHR = adjusted Hazard Ratio, CI = Confidence Interval

Among <80 years old persons, the hazard mitigation associated with vaccination was independent of the variant. Compared to those with primary vaccination, unvaccinated had a 10·8 (95% CI: 5·1-22·9) higher risk in 18-39 years, 6·9 higher risk (95% CI: 5·3-9·0) in 40-64 years and 2·2 (95% CI: 1·7-2·7) in 65-79 years age groups. In those three age groups, no additional reduction in risk was associated with the booster dose. In people > 80 years of age, the risk was divided by 3·7 (95% CI: 1·5-9·1) with a booster, compared to primary vaccination in the Omicron group [aHR=0·27; 95% CI: 0·11-0·65]. In the Delta group, there was no evidence of additional protection with the booster [aHR=0·82; 95% CI: 0·48-1·40]. In a supplementary analysis, the primary vaccination and booster modalities were split in two according to the median time between the booster dose and the virological diagnosis. This analysis did not show a relationship between this time and hazard reduction.

## Discussion

### Key results

Our national cohort study confirms a lower severity of Omicron infections compared to Delta infections in adults, by quantifying the risk of a severe hospital event, i.e. admission in ICU or death during hospitalisation. We defined this outcome as death may occur in patients not admitted to ICU wards, such as frail elderly patients^12^. People aged 40 to 64 years had a 9·1 times lower risk of severe outcome when infected with the Omicron variant (aHR=0·11). In the 80 years old and above the risk associated with Omicron was only half than that associated with Delta (aHR=0·51, p=0.052).

Substantial decrease in severity of Omicron cases compared with Delta cases has been described in recent studies. In South Africa, after controlling for factors associated with hospitalisation, individuals with S Gene Target Failure (SGTF), used as a proxy for Omicron infection, had lower odds of being admitted to hospital compared to non-SGTF infections, used as a proxy for Delta infection (0·2, 95% CI: 0·1-0·3); however, the Delta and Omicron cohorts were not contemporaneous and the Omicron cohort had more acquired immunity, which might have lessened the estimated severity of Omicron.^13^ Studies with contemporary cohorts have since been conducted in Canada,^14^ USA,^15^ UK^16^ and Norway.^17^ Our method and results align with the Canadian study; this was a matched cohort design, which described an aHR of 0·17 (95% CI: 0·08-0·37) in admission to ICU or death for Omicron cases when compared to Delta cases. In the USA (California), the HRs between Delta and Omicron cases, after a positive outpatient test, were 0·26 (95% CI: 0·10-0·73) for admission to ICU and 0.09 (95% CI: 0·01-0·75) for death. In the UK, the aHR was 0·41 (95% CI: 0·39–0·43) for hospital admission and 0·31 (95% CI: 0·26–0·37) for death. In Norway the HRs for hospitalisation between Delta and Omicron cases was 0·27 (95% CI: 0·20–0·36).

Our study also quantified the protection provided by vaccination. In Norway, an interaction between the variant and vaccination status was detected in the subgroup of people with primary vaccination completed 7-179 days ago, suggesting a weaker effectiveness of the vaccine for Omicron cases than for Delta cases (66% and 93%, respectively).^17^ We did not find evidence of such an interaction, except for the over-80s, although the size of our cohort is larger. However, our study focused on severe hospital events whereas the Norwegian study considered all hospital admissions, suggesting that the effectiveness of vaccination against the most severe forms of COVID-19 may be equivalent for both variants.

If initial *in-vitro* studies showed drastic reduction in neutralization of Omicron by sera from vaccinated individuals;^18^ epidemiological investigations later showed that vaccination and previous infections still conferred some levels of protection against Omicron, especially against severe diseases.^2,19^ Cellular immunity, especially T-cell response, has been shown to largely cross-react with Omicron and protect against severe COVID-19, even in cases with low levels of neutralizing antibodies.^20,21^ The large proportion of epitopes conserved between Omicron and previous variants, especially non-Spike epitopes, seems to be an important driver of T-cell protection against Omicron.^21,22^ A modelling study from South Africa estimated that severe COVID-19 outcomes during the Omicron wave were mostly reduced due to prior infection and/or vaccination.^11^ However, intrinsically reduced virulence still accounted for 25% of the risk reduction compared to Delta. Animal model studies also showed milder clinical pictures in Omicron-infected animals compared to Delta, which was associated with lower viral load in lower airways.^23,24^ Differences in the tropism of Omicron, which has been shown to rapidly infect bronchial cells but less efficiently infect pulmonary cells compared to Delta, might explain these mild symptoms.^25,26^ Therefore, the milder clinical presentation associated with Omicron seems to rely on both molecular specifics of the virus and immunological characteristics of the population.

Our study also showed evidence of age and variant-dependence in the magnitude of the risk reduction following a booster vaccination, relative to primary vaccination alone. While a statistically significant reduced risk of a severe hospital event was observed among the 80+ years old population infected with Omicron (aHR= 0·27, 95% CI: 0·11-0·65, p=0·003), this was not found in the younger age groups and in Delta cases of the same age group. In the UK, booster vaccination with an mRNA vaccine has been shown to be highly protective against hospitalisation and death in Omicron cases; however, the additional protection provided by the booster, compared to primary vaccination, was not quantified.^16^ This limited additional protection of the booster may be attributed to the level of residual protection against severe disease provided by primary vaccination. As mentioned earlier, research studies have demonstrated intact cellular immune response (SARS-CoV-2 spike-specific CD4+ and CD8+ T cells) elicited by vaccination against Omicron.^29-32^ Though this may be true for the younger population a diminished cellular immune response may be seen in the older population.^27^

It should be pointed out that, as the individuals were symptomatic at inclusion, our results did not take into account either the possible differential capacity of the two variants to cause symptomatic infection, or the capacity of the vaccines to prevent symptomatic infection. Further studies are needed to better understand the differences between Delta and Omicron ahead of symptomatic infection. The fact that we observed 44% of unvaccinated individuals in the Delta group and only 20% in the Omicron group supports a partial vaccine escape for the prevention of symptomatic infection by the Omicron variant as reported in other studies.

The risk of ICU admission or death was almost three times higher for patients with comorbidities than for those without, with a tendency to be higher for very-high-risk comorbidities. The absence of interaction showed that this excess risk was present for both variants. The higher risk of developing severe COVID-19 infection in patients with underlying medical comorbidities has been widely described and may lead to up to a six-fold increase in hospitalisation.^28^

The risk of ICU admission or death was almost two times higher among males compared to females as reported in other studies.^28-30^ The absence of interaction evidenced in our study is in favor of this excess risk existing for both variants.

### Strengths of the study

Studies comparing consecutive waves of the epidemic may lead to confounding biases; the decrease in disease severity in the Omicron cases could reflect an increase in immunity, whether natural or acquired through vaccination, a change in the characteristics of the infected population or a change in hospital care over time. In our study, to control for this bias, each Omicron case was matched according to age and date of diagnosis to a Delta case. In addition, a multivariate analysis was used to adjust the HR of severe events between Delta and Omicron variants infections on gender, age, vaccination status and comorbidities, which are known risk factors for severe forms of COVID-19. Cox proportional hazard model allowed for different follow-up times.

All cases included in this study were screened by RT-PCR in order to identify a set of target mutations. We designed an algorithm to classify cases in suspected Delta cases, suspected Omicron cases and other variant/inconclusive. Using cases for which a sequencing result was also available, this algorithm showed a diagnostic specificity of 99·1% for Delta and 92·9% for Omicron. PCR screening did not allow differentiate between the BA.1 and BA.2 Omicron sub-lineages, but as of January 15, 2022, BA.2 was very rarely detected in France.

Our study focused on severe hospital events, and only those stated “for Covid” were retained. In addition, only symptomatic cases were included. These three points may lead to a more specific signal and explain why we observed a higher risk reduction for Omicron cases compared to Delta cases, than other studies considering all RT-PCR cases and all hospital admissions, regardless of severity and imputability.

### Limitations

This study was designed to compare the severity of the Delta and Omicron variants, not to assess vaccine effectiveness. Therefore, further studies are needed to confirm the result we observed in those who received a booster compared to the primary vaccinated in different age groups. Although, the status of a known previous infection is used to define the required number of doses administrated (e.g. one single dose for primary vaccination in case of known previous infection), it was not possible to account for history of previous infection in our model.

Another limitation is the imperfect merge, by pseudonym, between the three databases: 16% of the persons fulfilling the inclusion criteria in SI-DEP were excluded because they were not found in VAC-SI. It is also possible that not all hospital events were found in SI-VIC. In addition, deaths which occurred elsewhere than in hospital were not taken into account. However, there is no reason why this loss of information should differ between the two groups of the cohort and therefore affect the hazard ratios.

## Conclusion

This cohort study confirms the lower severity of Omicron compared to the Delta variant, which has been suggested since the emergence of this variant. The protection offered by the vaccines does not appear to differ between the two variants, except in the over 80s. The vaccine protection is critical in the elderly as they have a higher risk of severe hospital events following infection with Omicron than younger subjects, even if this risk is much lower than with Delta.

## Supporting information

Supplemental 1 - Mutation screening

Supplemental 2 - All Hospital admission

## Data Availability

While all data used in this analysis were pseudonymised, the individual-level nature of the data used risks individuals being identified, or being able to self-identify, if it is released publicly. Requests for access to the underlying source data should be directed to Public Health France and will be granted in accordance with the GDPR and French Law.

## Authors contributions

SV, DLB, IPC, BC and YLS conceived and designed the study; SV, JS an LF reviewed the literature; JS, BC, and LF designed the algorithm for assigning a variant using PCR screening results which specificity was assessed by JS and VA; CM and VA managed the datasets. VA did the statistical analysis with the help of YLS and LF. ; VA and SV wrote the original draft, and all authors reviewed and edited the manuscript.

## Acknowledgments

We would like to thank the health care professionals and administrative teams in hospitals and biomedical laboratories, the professionals in charge of vaccinations and all those who contributed to the data management and analysis over the past two years. Our thoughts are also with all those who have been affected by the disease, and who are not just *cases*.

## Statements

### Data sharing

While all data used in this analysis were pseudonymised, the individual-level nature of the data used risks individuals being identified, or being able to self-identify, if it is released publicly. Requests for access to the underlying source data should be directed to Santé publique France and will be granted in accordance with the GDPR and French Law.

### Ethical statement

This study did not involve the human person. It was carried out by Santé Publique France using data collected by the Ministry of Health to manage the COVID-19 crisis. This processing of personal pseudonymised data was implemented in accordance with the legislative and regulatory prerogatives granted to Santé publique France to fulfil its public interest mission and in compliance with the provisions of the GDPR. In this context, the opinion of an ethics committee was not required.

