## Supplemental 1 - Mutation screening for "Severe hospital events following symptomatic infection with Sars-CoV-2 Omicron and Delta variants in France, December 2021 – January 2022: a retrospective, population-based, matched cohort study"

### Definition and validation of an algorithm using mutation screening results to classify SARS-CoV-2 cases as Delta or Omicron

Justine Schaeffer, Lucie Fournier, Vincent Auvigne

Santé publique France - 12 rue du Val d'Osne - 94415 Saint-Maurice - France

#### PCR screening strategy in France

In France, a proportion of samples which tested positive for SARS-CoV-2 by reserve transcription followed by quantitative PCR (RT-qPCR) are screened for specific mutations. The mutations included in the PCR screening strategy are chosen due to their potential or demonstrated impact on SARS-CoV-2 transmissibility, severity or immune escape. Such screening results are not sufficient to confirm an infection by a given variant, however, based on the variants circulating at a given time, some specific mutation profiles can allow to suspect a given variant. From 30 May 2021 on, a nation-wide PCR screening strategy was defined, targeting S mutations E484K, E484Q and L452R. Screening results are submitted to the national SARS-CoV-2 test database in three variables named A (for E484K), B (for E484Q) and C (for L452R). For these three variables, 0 indicates that the target mutation was not detected, 1 that it was detected, 8 that the result was inconclusive and 9 that the target mutation was not tested.

Following the emergence of Omicron in late November 2021, the French PCR screening strategy was adjusted. As over 99% of Omicron sequences did not carry any of the three mutations targeted by the PCR screening, a close follow-up of the A0B0C mutation profile (absence of E484K, E484Q and L452R) was initiated. However, this A0B0C0 profile was not specific of Omicron, with other circulating variants such as B.1.640 corresponding to this profile. Therefore, a new variable, called D, was added to national database. This variable D combined several mutations which were more specifically associated with Omicron, a D1 result indicating that one or several of these mutations were tested and found and a D0 result that one or several of these mutations were tested and not found. Between 29 November and 19 December 2021, D included S deletion 69-70 (DEL69-70) as well as mutations K417N and N501Y. Since 20 December 2021, D includes S DEL69-70 as well as mutations K417N, S371L-S373P and Q493R. Since 20 December 2021, E484Q (code B) is no longer tested and the follow-up of A0B0C0 has been shifted to A0C0.Table 1 : Evolution of the French PCR screening strategy over time. Columns "Mutation" and "Code" indicate target mutations and the database variable associated with each mutation. Known effects of each mutations on SARS-CoV-2 transmissibility and immune escape and the main variants carrying these mutations are also indicated.

| Time frame | Code | Mutation | Transmission | Immune escape | Variant |
|---|---|---|---|---|---|
| 30/05/2021<br>29/11/2021 | A | E484K | | + | Beta/Gamma |
| | B | E484Q | | + | some Alpha/Delta |
| | C | L452R | + | + | Delta |
| 29/11/2021<br><br>20/12/2021 | A | E484K | | + | Beta/Gamma |
| | B | E484Q | | + | some Alpha/Delta |
| | C | L452R | + | + | Delta |
| | D | DEL69-70<br>and/or K417N<br>and/or N501Y | + | +?<br>+? | Omicron<br>Omicron<br>Omicron/B.1.640 |
| 20/12/2021<br>… | A | E484K | | + | Beta/Gamma |
| | C | L452R | + | + | Delta |
| | D | DEL69-70<br>and/or K417N | | +?<br>+? | Omicron<br>Omicron |

| | | |
|---|---|---|
| | and/or S371L-S373P | Omicron |
| | and/or Q493R | Omicron |

#### Algorithm definition

Over the study period (01/12/2021 – 10/01/2022), the main variants detected by random sequencing in metropolitan France were Delta (79.1% of 22 493 interpretable sequences), Omicron (20.5%) and B.1.640 (0.4%), with additional variants being detected sporadically. S mutation L452R (coded C1) is mainly found in Delta variant. Among the mutations included in the D variable (code D1), S371L-S373P and Q493R are found almost exclusively in Omicron. DEL69-70, K417N and N501Y can be found in several variants, but over the study period, they were mostly associated with Omicron. The addition of D in the PCR screening strategy aimed at following-up more specifically Omicron, but it was progressively implemented over the study period and is not always available. Mutations profile A0C0 (absence of E484K and L452R) could correspond to Omicron but also to other variants (B.1.640, B.1.1.318, etc…). However, as these variants were much less frequent than Omicron over the study period and at national scale, A0C0 was used in addition to D1 as a proxy for Omicron.

The following algorithm was developed to suspect infection by a variant (Omicron, Delta or other) using PCR screening results:

- Suspected Delta case
  - L452R found (C1)
  - **AND** mutations associated with Omicron not found (D0) or not available (D=NA)
- Suspected Omicron case
  - Mutations associated with Omicron found (D1) or not tested (D9) or inconclusive (D8)
    **AND** E484K not found (A0)
    **AND** L452R not found (C0)
  - **OR** mutations associated with Omicron not available (D=NA)
    **AND** E484K not found (A0)
    **AND** L452R not found (C0)
    **AND** E484Q not found (B0), not tested (B9) or not available (B=NA)
- Other variant or inconclusive for all other cases

#### Algorithm validation

To assess the performance of the above-mentioned algorithm to assign a variant using PCR screening results, its results were compared with variants identified by sequencing. Among the 850 824 cases included in the study, 13 084 cases had both a PCR screening result and a sequencing result. Sequencing result was inconclusive for 1 510 cases, which were excluded from the analysis. As the nomenclature for sequencing results in the database was not homogeneous, the variant identified by sequencing was defined as follow:

- Delta variant if the sequencing result included
  - Delta
  - **AND/OR** 21A or 21I or 21J (Clade)
  - **AND/OR** B.1.617.2 or AY. (Lineage)
- Omicron variant if the sequencing result included
  - Omicron
  - **AND/OR** 21K or 21L or 21M (Clade)
  - **AND/OR** B.1.1.529 or BA. (Lineage)
- Other for every other case

The comparison between the suspected variant obtained using the PCR screening algorithm and the confirmed variant obtained by sequencing are presented in table 3.

Supplementary material to « Severe hospital events following symptomatic infection with Sars-CoV-2 Omicron and Delta variants in France, December 2021 – January 2022: a retrospective, population-based, matched cohort study »

*Table 2: comparison of algorithm results (based on PCR screening) and sequencing results*

| | | Sequencing | | | |
|---|---|---|---|---|---|
| | | OTHER | DELTA | OMICRON | Total |
| PCR screening | OTHER or inconclusive | 41 | 265 | 254 | 560 |
| | DELTA | 12 | 6370 | 31 | 6413 |
| | OMICRON | 153 | 360 | 4088 | 4601 |
| | Total | 206 | 6995 | 4373 | 11574 |

| | OTHER | DELTA | OMICRON |
|---|---|---|---|
| Sensitivity | 19,9% | 91,1% | 93,5% |
| Specificity | 95,4% | 99,1% | 92,9% |
| PPV | 7,3% | 99,3% | 88,9% |
| NPV | 98,5% | 87,9% | 95,9% |
