## Supplemental 2 - All Hospital admission for "Severe hospital events following symptomatic infection with Sars-CoV-2 Omicron and Delta variants in France, December 2021 – January 2022: a retrospective, population-based, matched cohort study"

### Methods

In our main paper[1], the outcome is the occurrence of a severe hospital event. In this supplement, the results are presented with hospital admissions as the outcome. The event is the first hospital admissions following virological diagnosis (or the day of diagnosis), regardless of the type of ward. Apart from this definition of the outcome, all methods are those of the main paper.

### Results

*Table 1. Hospital admissions observed during the follow-up period, December 2021-January 2022, France, (n= 184 364)*

| Characteristic | DELTA, N = 92 182 | OMICRON, N = 92 182 | p-value |
|---|---|---|---|
| Hospital admissions | 1 845 (2·0%) | 371 (0·4%) | <0.001 |

[1] Severe hospital events following symptomatic infection with Sars-CoV-2 Omicron and Delta variants in France, December 2021 – January 2022: a retrospective, population-based, matched cohort study

*Figure 1. Evolution since the virological diagnosis of the cumulative probability of hospital admissions among cases infected by the Omicron or Delta variant of the SARS-CoV-2, December 2021-January 2022, France, (n= 184 364)*

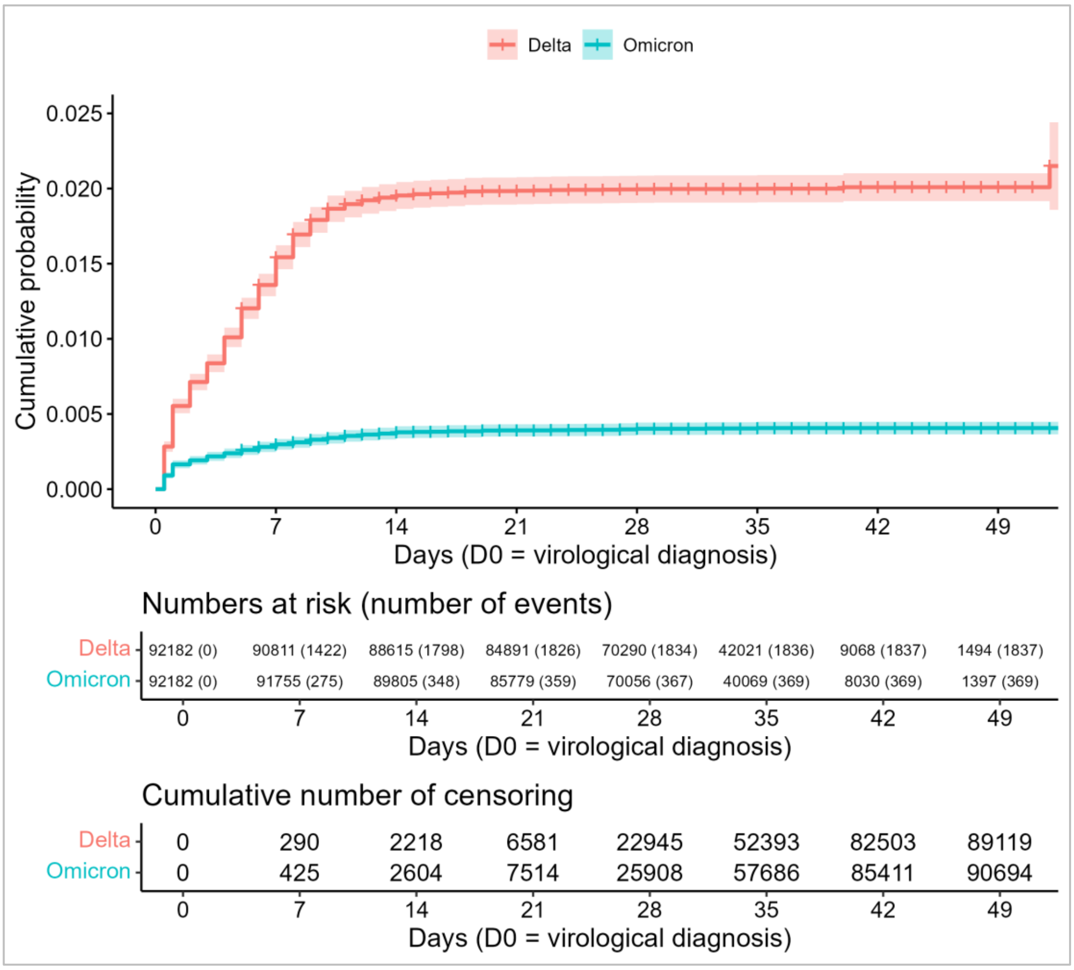


In the DELTA group of the cohort, 87% of hospital admissions were observed within 10 days of positive sampling, and 92% within 20 days. In the OMICRON group, 84% of events occurred within 10 days and 96% within 20 days (Figure 1).

*Table 2. Univariate analysis for the risk of hospital admissions among cases infected by the Omicron or Delta variant of the SARS-CoV-2, December 2021-January 2022, France (n= 184 364)*

| | | cHR (95% CI[1]) | p-value |
|---|---|---|---|
| Variant | DELTA | Ref | - |
| | OMICRON | 0.20 [0.18 to 0.22] | <0.001 |
| Age | [18,40) | 0.19 [0.16 to 0.22] | <0.001 |
| | [40,65) | Ref | - |
| | [65,80) | 5.61 [5.07 to 6.21] | <0.001 |
| | [80,Inf) | 15.31 [13.69 to 17.12] | <0.001 |
| Sex | F | Ref | - |
| | M | 1.6 [1.47 to 1.74] | <0.001 |
| Vaccination | Unvaccinated | 6.1 [5.52 to 6.74] | <0.001 |
| | Primary vaccination | Ref | - |
| | Booster | 2.28 [1.97 to 2.65] | <0.001 |
| Comorbidity | None | Ref | - |
| | Medium-risk | 5.38 [4.91 to 5.9] | <0.001 |
| | Very-high-risk | 9.07 [8.01 to 10.27] | <0.001 |

*[1]cHR = crude Hazard Ratio, CI = Confidence Interval (univariate Cox proportional hazard model)l*

*Table 3. Cumulative probability at D21 of hospital admissions among cases infected by the Omicron or Delta variant of the SARS-CoV-2, stratified by co-variables, December 2021-January 2022, France, (n= 184 364)*

| | | DELTA | | OMICRON | |
|---|---|---|---|---|---|
| Age | [18,40) | 355 | [297 - 413] | 57 | [34 - 80] |
| | [40,65) | 1,808 | [1645 - 1972] | 286 | [218 - 354] |
| | [65,80) | 8,035 | [7117 - 8943] | 1,812 | [1318 - 2303] |
| | [80,Inf) | 21,094 | [18253 - 23837] | 6,475 | [4717 - 8201] |
| Sex | F | 1,215 | [1105 - 1325] | 235 | [186 - 284] |
| | M | 1,919 | [1768 - 2069] | 380 | [311 - 448] |
| Vaccination | Unvaccinated | 3,122 | [2901 - 3342] | 640 | [490 - 790] |
| | Primary vaccination | 596 | [523 - 669] | 178 | [140 - 216] |
| | Booster | 1,838 | [1423 - 2251] | 514 | [366 - 662] |
| Comorbidity | None | 936 | [859 - 1014] | 168 | [135 - 201] |
| | Medium-risk | 4,579 | [4133 - 5023] | 1,103 | [853 - 1352] |
| | Very-high-risk | 6,587 | [5491 - 7671] | 1,697 | [1073 - 2317] |

*Kaplan-Meier estimator and 95% CI*

Several interactions involving age and variant are identified by the multivariate Cox proportional hazards model. (Table 4).

*Table 4. Multivariate analysis using Cox regression for the risk of hospital admissions among cases infected by the Omicron or Delta variant of the SARS-CoV-2, stratified by age, December 2021-January 2022, France (n= 149,064)*

| | HR1 | 95% CI1 | p-value |
|---|---|---|---|
| **Variant** | | | |
| DELTA | Ref | | |
| OMICRON | 0·23 | 0·17 to 0·30 | <0·001 |
| **Age** | | | |
| [18,40) | 0·29 | 0·20 to 0·42 | <0·001 |
| [40,65) | Ref | | |
| [65,80) | 9·11 | 7·00 to 11·9 | <0·001 |
| [80,Inf) | 41·8 | 31·0 to 56·5 | <0·001 |
| **Sex** | | | |
| Female | Ref | | |
| Male | 2·03 | 1·76 to 2·34 | <0·001 |
| **Vaccination** | | | |
| Primary vaccination | Ref | | |
| Unvaccinated | 6·53 | 5·48 to 7·79 | <0·001 |
| Booster | 1·49 | 1·08 to 2·07 | 0·016 |
| **Comorbidity** | | | |
| None | Ref | | |
| Medium-risk | 2·68 | 2·29 to 3·13 | <0·001 |
| Very-high-risk | 4·13 | 3·28 to 5·20 | <0·001 |
| **Variant * Age** | | | |
| OMICRON * [18,40) | 1·32 | 0·86 to 2·03 | 0·21 |
| OMICRON * [65,80) | 1·32 | 0·96 to 1·81 | 0·089 |
| OMICRON * [80,Inf) | 2·26 | 1·64 to 3·10 | <0·001 |
| **Variant * Vaccination** | | | |
| OMICRON * Unvaccinated | 0·64 | 0·49 to 0·84 | 0·001 |
| OMICRON * Booster | 0·63 | 0·45 to 0·86 | 0·004 |
| **Variant * Comorbidity** | | | |
| OMICRON * Medium-risk | 1·31 | 1·01 to 1·70 | 0·041 |
| OMICRON * Very-high-risk | 1·61 | 1·15 to 2·27 | 0·006 |
| **Age * Sex** | | | |
| [18,40) * M | 0·56 | 0·41 to 0·75 | <0·001 |
| [65,80) * M | 0·93 | 0·75 to 1·15 | 0·49 |
| [80,Inf) * M | 0·75 | 0·59 to 0·94 | 0·014 |
| **Age * Vaccination** | | | |
| [18,40) * Unvaccinated | 0·89 | 0·62 to 1·28 | 0·54 |
| [65,80) * Unvaccinated | 0·53 | 0·41 to 0·68 | <0·001 |
| [80,Inf) * Unvaccinated | 0·34 | 0·25 to 0·44 | <0·001 |
| [18,40) * Booster | 2·48 | 1·29 to 4·76 | 0·006 |
| [65,80) * Booster | 0·50 | 0·33 to 0·74 | <0·001 |
| [80,Inf) * Booster | 0·36 | 0·24 to 0·55 | <0·001 |
| **Age * Comorbidity** | | | |
| [18,40) * Medium-risk | 0·91 | 0·62 to 1·34 | 0·64 |
| [65,80) * Medium-risk | 0·68 | 0·54 to 0·85 | <0·001 |
| [80,Inf) * Medium-risk | 0·50 | 0·39 to 0·64 | <0·001 |
| [18,40) * Very-high-risk | 1·47 | 0·85 to 2·55 | 0·17 |
| [65,80) * Very-high-risk | 0·70 | 0·51 to 0·96 | 0·025 |
| [80,Inf) * Very-high-risk | 0·33 | 0·23 to 0·47 | <0·001 |

1HR = Hazard Ratio, CI = Confidence Interval
